# Spectral analysis of the daily evolution of deaths due to Covid-19 in France and in the world shows a weekend effect: myth or reality?

**DOI:** 10.1101/2020.06.23.20135442

**Authors:** André Constantinesco, Vincent Israel-Jost, Philippe Choquet

## Abstract

**Background:** The weekend effect has been extensively observed for different diseases and countries and recognized as a fact but without obvious causes.

**Objectives:** In this paper we first aimed at investigating the existence of a periodicity in the death count due to Covid-19, and second, at opening the discussion concerning the reality of this effect in this particular context.

**Methods:** Daily statistics of deaths due to the Covid-19 pandemic were subjected to a discrete Fourier transform spectral analysis for France and the world, over the periods from March 29 to May 16, 2020 and from January 22 to May 18, 2020 respectively.

**Results:** In both cases, a frequency peak of one harmonic corresponding to a period of 7.11 days was observed for France and the world. In France, this weekly frequency corresponds to a decrease in deaths every Sunday, whereas for the world the systematic decrease is shifted on average by 1.5 days and corresponds to Saturday or Friday.

**Conclusion:** At the world scale and for the epidemic period we confirm the existence of a consecutive weekend effect in the context of the Covid-19 pandemic.

## Introduction

The weekend effect^1-4^ has been described as a periodic pattern among the relative numbers of deaths per year linked with admission on weekends, for many diseases but its causes are still debated.

Since its emergence, the Covid-19 pandemic has resulted in a significant number of deaths every day in France and the rest of the world. The daily recording of death provides daily data taken country by country and then counted for all of them, giving a so-called global view (at least for the affected countries where these data are recorded and accessible). For one month of observation a weekly periodicity of deaths and daily contaminations due to Covid-19 had already been noted in May 2020 in 7 countries out of the 12 analyzed^3^ and interpreted as intergenerational interactions that may be at the origin of this periodicity. In order to confirm the existence of this week periodicity on a large scale (world data and large observation period), we propose to search for periodic events in the daily evolution of the data accounting for deaths due to Covid-19 using a discrete Fourier transform spectral analysis (method applicable to any other pandemic or epidemic). Contrary to the study previously mentioned, a spectral analysis has the advantage of avoiding any modeling of the statistical data. Of course, the representative curves of this evolution show a main tendency: a daily increase in the number of deaths during the first weeks when the virus circulates, before slowly decreasing following the sanitary containment measures taken by the governments. But Fourier analysis reveals the subtler behavior of the curve within this main pattern and confirms the existence of a weekly periodicity.

## Methods

Daily statistical data on deaths were collected from the Center for Systems Science and Engineering at Johns Hopkins University (Baltimore, USA) and are available at: https://coronavirus.jhu.edu/map.html. The spectral analysis of the data was performed by discrete Fourier transform using pro Fit software (version 7.0, Quansoft, http://www.quansoft.com/).

The representative curves of the daily variation of deaths (figure 1) are assimilated to positive and increasing real functions sampled every 24 hours and windowed by the total observation time, which is several months here. In this case, sampling is carried out at a frequency fe = 1/86400 Hz corresponding to a sampling period Te of 24 hours. The recording of the N samples lasts a time T= N.Te = N/fe corresponding to the analysis time window, i.e. 49 and 118 days for France and the world respectively.

**Figure 1:**
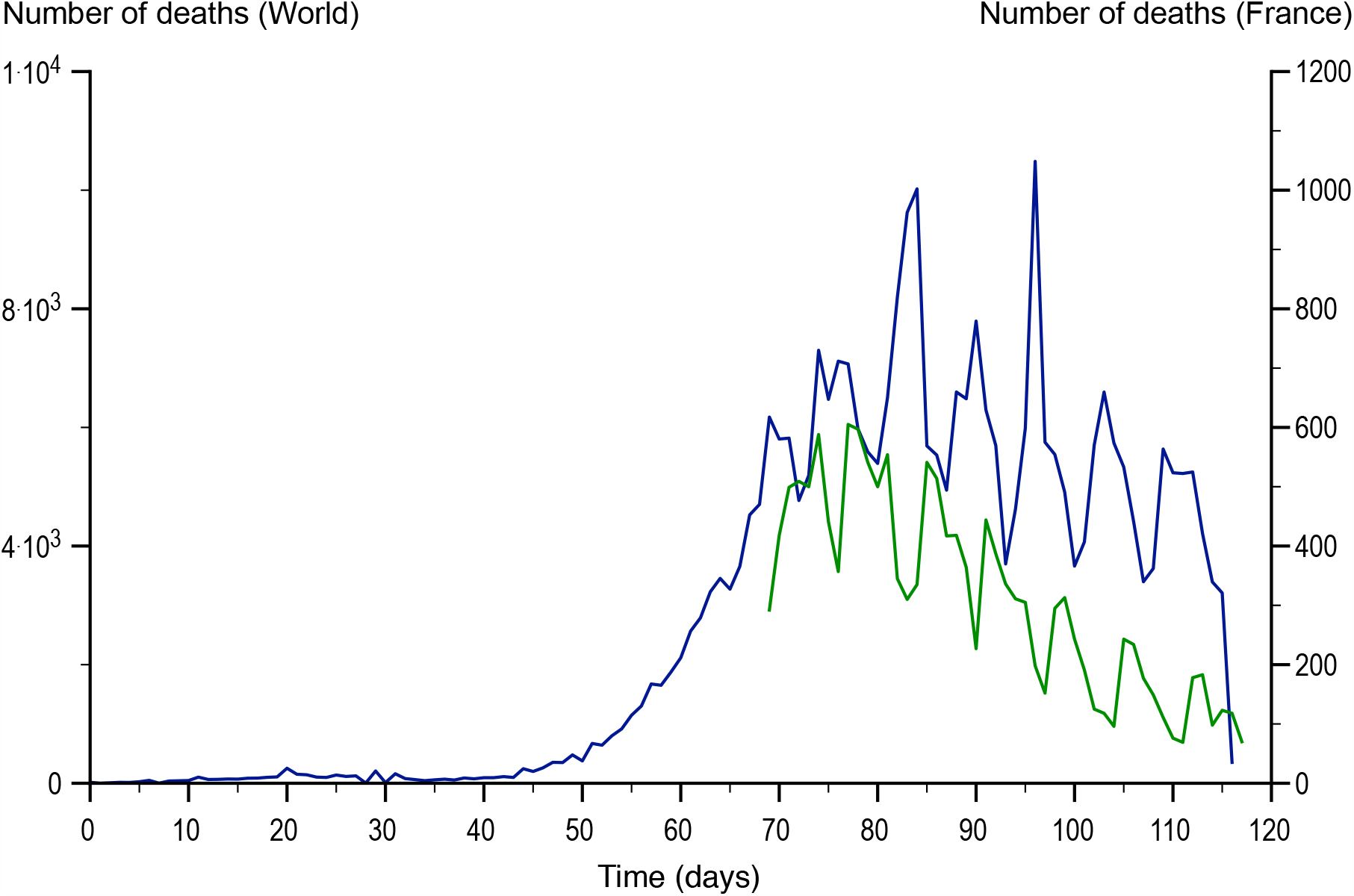
Daily evolution of mortality due to Covid-19, in the world (blue curve) and in France (green curve) where the pandemic appeared on the 69th day compared to the beginning of the recording of global data.

From these N samples, the discrete Fourier transform is used to calculate N points of the spectrum defined by their abscissa f(k) and their ordinate S(k). Thus the abscissa of each point of the spectrum are:

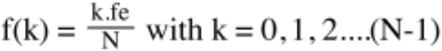

and the ordinates of the points on the spectrum are:

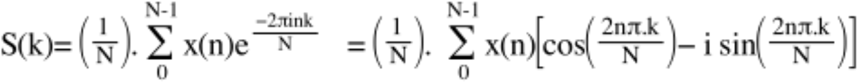

## Results

The spectral analysis of the two functions representing the daily evolution of deaths in France and in the world highlights a first harmonic whose peak frequency is located at 1.627.10^−6^ Hz in both cases, figure 2. The period T_0_ of this harmonic corresponds to a duration equal to 7.11 days.

**Figure 2:**
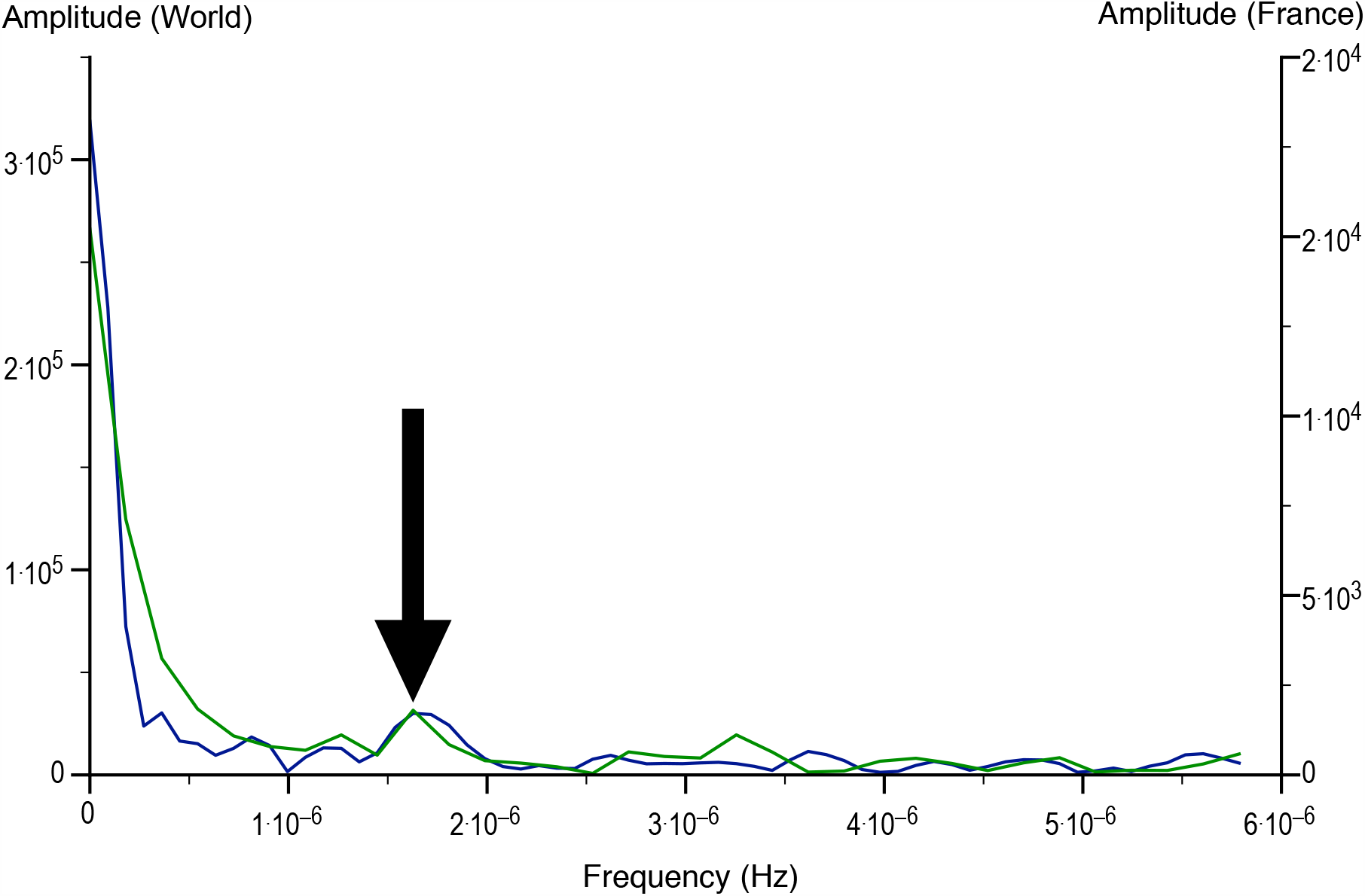
Spectral analysis of the curves shown in Figure 1, for the world (blue curve) and for France (green curve) highlighting (arrow) a harmonic centered at 1.6.10^−6^ Hz.

Figure 3 shows the curves of deaths in France and in the world when they are centered on the common window, starting on the date of the beginning of the French count (e.g. 04/29/2020). In this figure, one immediately observes a periodic decrease in mortality, weekly from Sunday to Sunday for France and centered on Fridays (4 times) or Saturdays (3 times) for the world (Table 1). This periodicity displays a mean of 7.14 days for the world and 7 days for France in accordance with the spectral analysis.

**Table 1.**

| **Death minima world** | **Death minima France** |
| --- | --- |
| 3/27 friday | 3/29 sunday |
| 4/03 friday | 4/05 sunday |
| 4/11 saturday | 4/12 sunday |
| 4/18 saturday | 4/19 sunday |
| 4/24 friday | 4/26 sunday |
| 5/08 friday | 5/10 sunday |
| 5/16 saturday |  |
| Mean: 7·14 days | Mean: 7 days |

## Discussion

Comparing our results with those presented by I. Ricon-Becker and colleagues^5^, we confirm the existence of a weekly periodicity of daily deaths but for the whole world and not only in some countries. Then we observed that the systematic decrease in daily deaths is at the end of the week in the world (Friday-Saturday) and every Sunday in France. Finally, compared to spectral analysis, which makes no modeling hypothesis, the methodology of I. Ricon-Becker et al.^5^ uses data smoothing with a 3-day moving average followed by a statistical analysis based on autocorrelation before applying a sinusoidal model in which 4 parameters are fitted. Spectral analysis is simpler and more robust for accurately determining a periodicity in a time data set.

We will consider three main hypotheses likely to be at the origins of this weekly cycle corresponding to a minimum of deaths, in France as in the world, on weekends or even Sundays.

The three possibilities are:

1. Couting. The observed weekend effect could follow from difficulties to ensure homogeneous counting of deaths during weekends.
2. Societal effects that are likely to punctuate the daily evolution of mortality or
3. A biomedical effect linked to hospital structures and the means of patient care.

1. One possibility that cannot be excluded at first, would be that the minimum number of deaths on weekends is simply linked to the fact that data may not be transmitted everywhere on weekends, a hypothesis already formulated to explain the weekend effect.^6^ Indeed, it is impossible to know whether the daily sampling of mortality data is carried out strictly on a daily basis, given that it is certainly the result of different collection and transmission arrangements. The daily counting of Covid-19 deaths is delicate because the collection of data in real time is patchy and the methods vary from one country to another. Thus, as the French national press^7-9^ and the international press^10,11^ regularly point out, Spain, Germany, Luxembourg and South Korea count all the deaths of people testing positive for Covid-19, whether in hospital or outside. In Belgium, where nursing homes officially concentrate more than half of the fatal cases related to this disease, the figures include untested victims of the coronavirus but suspected to have been affected. The French figures also include deaths in retirement homes. In China and Iran only deaths in hospitals are counted, and in the United States the accounting of deaths varies from state to state. The Covid-19 weekend effect may also be due to the fact that the counting of weekend deaths is delayed to mondays or tuesdays but with the exact date of the death being also shifted for commodity reasons as it has been pointed out for France^12^, which could explain the French weekend effect we measured. Finally the homogeneity of the deaths world data could also be influenced by the countries lamenting the most severe infection. However, given this complexity in the collection and transmission of statistics, one would expect for the pooled world data to be averaged rather than this almost weekly periodicity.
2. One can then ask questions such as I. Ricon-Becker et al.^5^ on a societal origin despite the cultural and religious differences existing between the countries that feed the Covid-19 global database of deaths. The observed rhythm of the 7-day week (circaseptan rhythm) is obviously not a rhythm of biological origin but a human construction organizing our lives. A mortality weekend effect in medicine is known^1-4^ and is the subject of articles and discussions without reaching a consensus on its causes.^4,13,14^ The observation on which this effect is based, shows an excess or a decrease of weekend deaths, depending on the diseases but also on many other factors including seasonality for instance.^15^ Several studies have shown that certain social phenomena can, on a global scale, be regulated by the week, such as the increase in the suicide rate^16^ or cardiovascular accidents^17^ on mondays, with the hypothesis of a link with the return to work after two days of rest. In the case of the Covid-19 pandemic, a societal cause that would lead to periodic deaths remains to be demonstrated. However, in this particular context, given that the average incubation period is between 11.5 and 15.6 days^18^ and that public transport as well as working places are locations of contamination, the regularity of weekends limiting travel and group work (even during periods of containment) could, for example, have a link with the weekly death rate. On the contrary even if the emergence and spread of this virus is not independent of the behaviour of human societies, the countries affected have, at different times, put in place barrier measures at different times (physical distancing, wearing of masks, etc.) as well as instructions for population containment and restrictions on social activities, but these have not been applied in the same way everywhere. Therefore, these prevention measures, which vary over time and by country, are more likely to smooth out daily mortality data rather than to favour periodicity.
3. Similarly, a possible influence of the reception and management of Covid-19 patients in intensive care units or specialized hospital structures, even if they have shown differences in approach, methodology and resources throughout the world, cannot logically be considered to have any influence on the deaths week cycle.^19,20^ Finally, the worldwide involvement of healthcare personnel in the fight against this pandemic, which has necessitated an urgent reorganization of the healthcare offer, does not allow for a decrease in the quality of care during weekends either. One limitation of this work, however, remains the reliability, which we cannot guarantee, of the statistical data used.

Therefore, it would be interesting to take daily statistical data from other pandemics that have affected the world and apply spectral analysis to see whether a periodicity, possibly similar, also emerges.

In this work we applied a discrete Fourier transform spectral analysis to the data representing the daily variations of Covid-19 deaths around the world and particularly in France. This methodology, which does not use any data modeling, has allowed us to highlight a weekly periodicity of deaths with a decrease in systematic mortality at the end of the week (Friday-Saturday) for the world and on Sunday for France. It should be noted that very few studies^21-23^ in epidemiology make use of spectral analysis in temporal terms.

Finally, and subject to the limits previously discuted, a consecutive week periodicity on the daily counting of deaths due to Covid-19 for the pooled world data is a mathematical reality but its causes remain at present as difficult to assess as the other weekend effects already described.

## Data Availability

Data are freely available on line.

https://coronavirus.jhu.edu/map.html

## Contributors

AC organized and entered data. AC, VIJ and PC contributed to data interpretation. AC drafted the manuscript. All authors critically revised the drafted manuscript and approve of the submitted manuscript.

